# Benchmark Pathology Report Text Corpus with Cancer Type Classification

**DOI:** 10.1101/2023.08.03.23293618

**Authors:** Jenna Kefeli, Nicholas Tatonetti

## Abstract

In cancer research, pathology report text is a largely un-tapped data source. Pathology reports are routinely generated, more nuanced than structured data, and contain added insight from pathologists. However, there are no publicly-available datasets for benchmarking report-based models. Two recent advances suggest the urgent need for a benchmark dataset. First, improved optical character recognition (OCR) techniques will make it possible to access older pathology reports in an automated way, increasing data available for analysis. Second, recent improvements in natural language processing (NLP) techniques using AI allow more accurate prediction of clinical targets from text. We apply state-of-the-art OCR and customized post-processing to publicly available report PDFs from The Cancer Genome Atlas, generating a machine-readable corpus of 9,523 reports. We perform a proof-of-principle cancer-type classification across 32 tissues, achieving 0.992 average AU-ROC. This dataset will be useful to researchers across specialties, including research clinicians, clinical trial investigators, and clinical NLP researchers.

## Introduction

Patient data derived from structured electronic health records or molecular sequencing are frequently used as input for clinical models across cancer types. However, unstructured free text, such as pathology reports or clinical notes, is less frequently used in biomedical data analysis, despite being regularly generated as part of the EHR. Tumor pathology reports in particular are an essential source of clinical data, often containing nuanced information that is not always captured within structured datasets. Report text generally includes a macroscopic description of tumor appearance, location, and size; microscopic description of tissue structure and cell differentiation; evaluation of margins; and sometimes genetic or immunohistochemistry results. Reports can also contain the patient’s stage and grade, which help inform treatment, clinical care management, and prognosis.

Despite the potential utility of pathology report data in clinical research efforts, there currently does not exist a large, de-identified public dataset of pathology report text. However, The Cancer Genome Atlas (TCGA) has made available over 11,000 de-identified PDF reports associated with patient samples in their repository.^1^ TCGA is a particularly rich data source, containing clinical metadata, tumor genomic data, histopathology slide images, and follow-up patient tracking for survival outcomes.

Although available for download, pathology reports from TCGA have not been extensively utilized for research purposes due to their PDF-formatting, highly varied structure, and the presence of image-artifacts, making automated analysis difficult.

Numerous recent studies have utilized information derived from pathology reports, both in general^2–13^ and using subsets of the TCGA pathology report dataset.^14–20^ Previous studies incorporating TCGA pathology report data have largely relied on manual curation and limited term-set extraction across smaller subsets of the dataset.

More recently, one study^21^ used OCR and NLP techniques on a TCGA subset of breast cancer patients for an information retrieval task. Another study combined OCR and traditional machine learning techniques to classify tumor grade using a TCGA subset of approximately 500 prostate cancer patients.^22^ Additionally, Allada et al. compared different NLP classification methods for the prediction of seven disease classes within a TCGA subset consisting of roughly 2,000 patients.^23^ The accelerated increase in recent research efforts using pathology report text as a basis for a variety of clinical prediction tasks demonstrates the utility of this type of data and the need for a benchmark dataset.

Here, we describe the curation of a text corpus derived from the set of all TCGA pathology reports. To convert pathology reports from PDF to machine-readable text, we employ OCR as well as significant OCR post-processing. After processing, translating from image to text, and cleaning, we leverage recent advances in NLP^24^ and its application to clinical text^25–27^ to demonstrate the utility of this dataset by training a cancer-type prediction model. We make the final corpus of 9,523 patient reports publicly available for researchers to use for data mining or machine learning applications. The corpus will be particularly useful for researchers who may not have access to institution-specific or otherwise controlled-access corpora and can potentially provide a benchmark in this field going forward.

## Results

### TCGA Pathology Report Pre-Processing and Data Selection

11,108 pathology reports, corresponding to 11,010 patients, were downloaded from the TCGA data portal. The dataset was pre-processed as follows: First, we removed 82 patients with multiple reports and 399 patients with non-primary tumors. Then, to ensure that the final dataset is fully complete with respect to associated outcome data, we removed 72 patients who did not have survival data in the TCGA Clinical Data Resource.^28^ This resulted in a selection of 10,457 patients, with each patient having one corresponding report, and all reports descriptive of primary tumors. Next, we removed 381 “Missing Pathology” reports, which were placeholder forms indicating a lack of pathology report for specific patients, as well as 14 reports of poor scan quality (see Methods). We additionally removed 212 “TCGA Pathologic Diagnosis Discrepancy Form” reports, which consisted mostly of diagnosis discrepancies indicative of inaccurate pathology reporting. After these filters were applied, 9,850 reports were processed through text extraction.

### Text Extraction and OCR Post-Processing

We used Optical Character Recognition (OCR) to transform reports from PDF into text. We qualitatively evaluated several different OCR programs, with Textract^29^ producing the most accurate results and being the best able to remove report artifacts (see Methods). We processed 9,850 reports (25,478 pages) through Textract, and then parsed and post-processed the resultant output files. Redaction bars and TCGA barcodes, artifacts of the TCGA quality control process, were removed by Textract (Fig S1A-B).

We removed reports that consisted partially or entirely of multiple-choice forms (see Methods). We identified and removed 210 “Consolidated Diagnostic Pathology Form” reports using a combination of keyword and selection element (check-box) filters. We also manually reviewed and removed any “Synoptic Translated” forms as well as any reports with a large amount of multiple-choice selection elements. After removal, 9,547 reports (24,214 pages) remained.

We additionally removed within-report TCGA metadata insertions, which occur at inconsistent coordinates and at varying angles. These insertions can interrupt sentences in the OCR translation and need to be removed for clean final text (Fig S1C). We removed quality control (QC) tables added to the reports by the TCGA, which contain information about sample quality but are irrelevant for diagnosis and not included in standard pathology reports. See Methods for full details on this process, but briefly, we used Textract to identify the QC table’s section headers and drew a custom bounding box around each detected term. We validated this approach on a prototype set of 50 randomly-selected reports, and found 100% concordance between manual and automatic identification of QC tables using our bounding box technique. We additionally utilized Textract’s word-level text-type annotation, removing lines that contain only hand-written words. These lines were added by TCGA during QC and were typically mis-translated by OCR; their removal improves the overall quality of the dataset. 24,099 pages remained.

Finally, to finish cleaning the dataset, we sought to remove clinically irrelevant and potentially confounding clinic-specific section headers from the remaining reports. To maintain the general utility of this dataset, we employed a conservative approach, only removing content that is clearly irrelevant to pathological description and patient diagnosis. We manually reviewed 500 randomly-selected report pages to compile a list of 312 regular expressions which were used to remove report lines (see Methods). More than 100,000 lines were removed in this step (Fig 1B).

**Figure 1.**
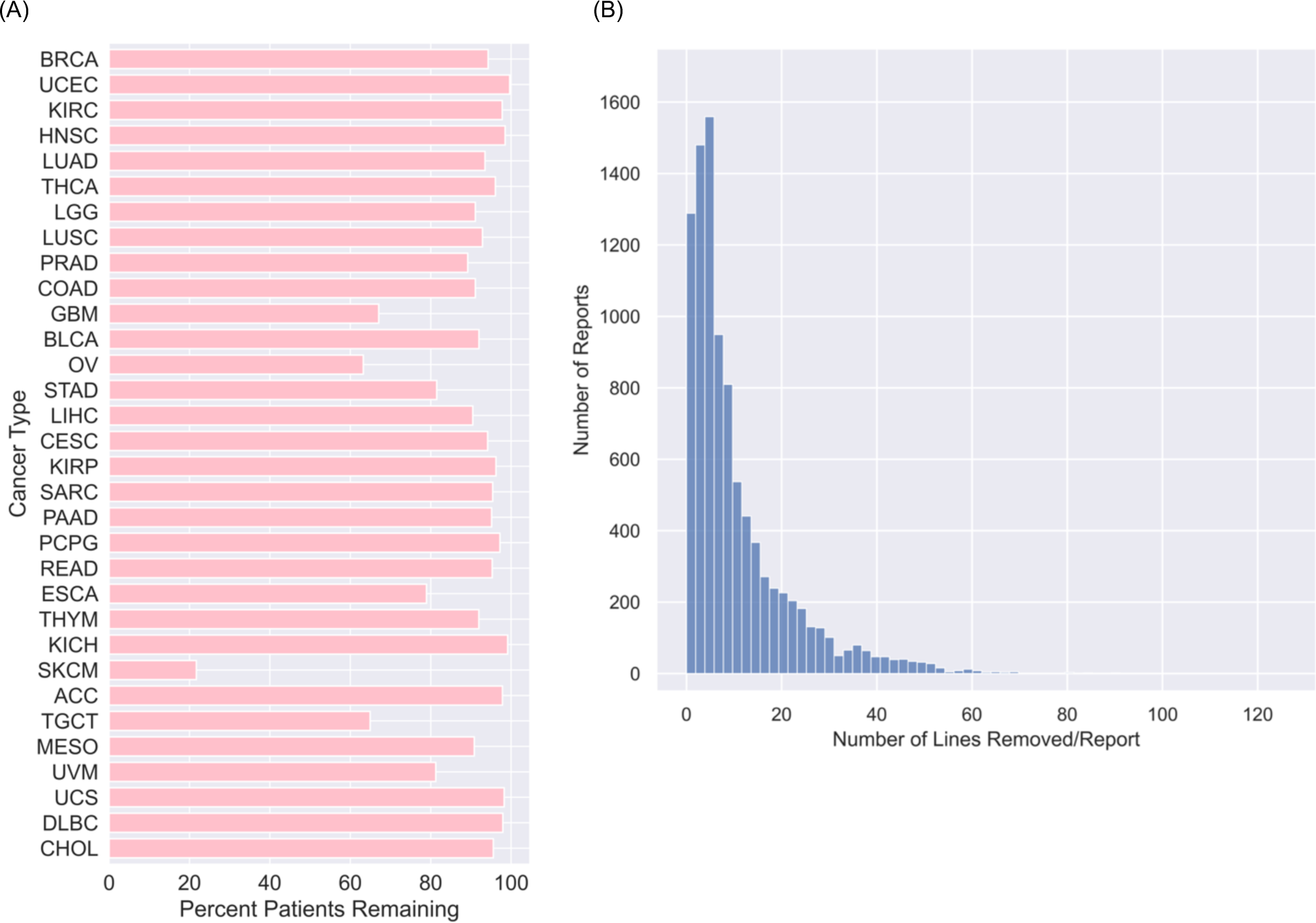
Patient and Line Distributions after Dataset Processing. (A) Distribution of patients remaining in the dataset after data selection, OCR, and post-processing, presented per cancer type. See also Table S1. (B) Distribution of number of lines removed per report during the final post-processing step of matched regular expression removal.

In total, 9,523 reports (23,909 pages, or 842,134 lines) remain in the final dataset (Fig 1A, 2A-D). The frequency of cancer type within the dataset varies: Breast invasive carcinoma is the most prevalent, with 1,034 patients, and Cholangiocarcinoma is the least prevalent, with 43 patients (Fig 2A). We compiled the demographic characteristics of the patient population in the final pathology report dataset overall (Table 1) and plotted distribution of demographics by cancer type (Fig S5).

**Figure 2.**
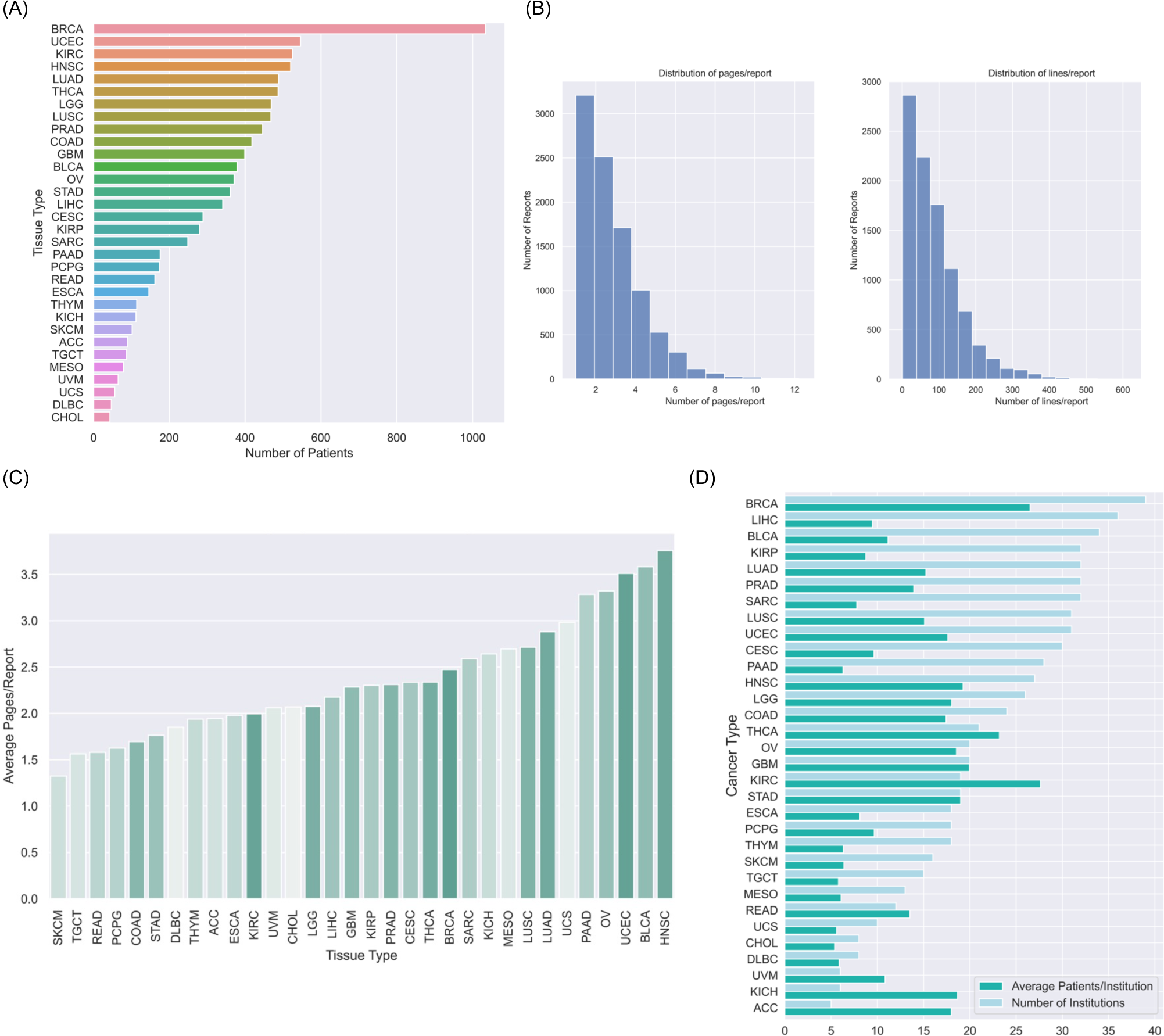
Final Dataset Characteristics. (A) Cancer type distribution, ordered by prevalence. (B) Distribution of number of pages per report (left) and lines per report (right). (C) Distribution of number of pages per report, segmented by tissue. Darker hue indicates greater prevalence of cancer type within this dataset. (D) Distribution of report- generating institutions (tissue source sites) and average number of reports per institution, presented separately by cancer type.

**Table 1.**
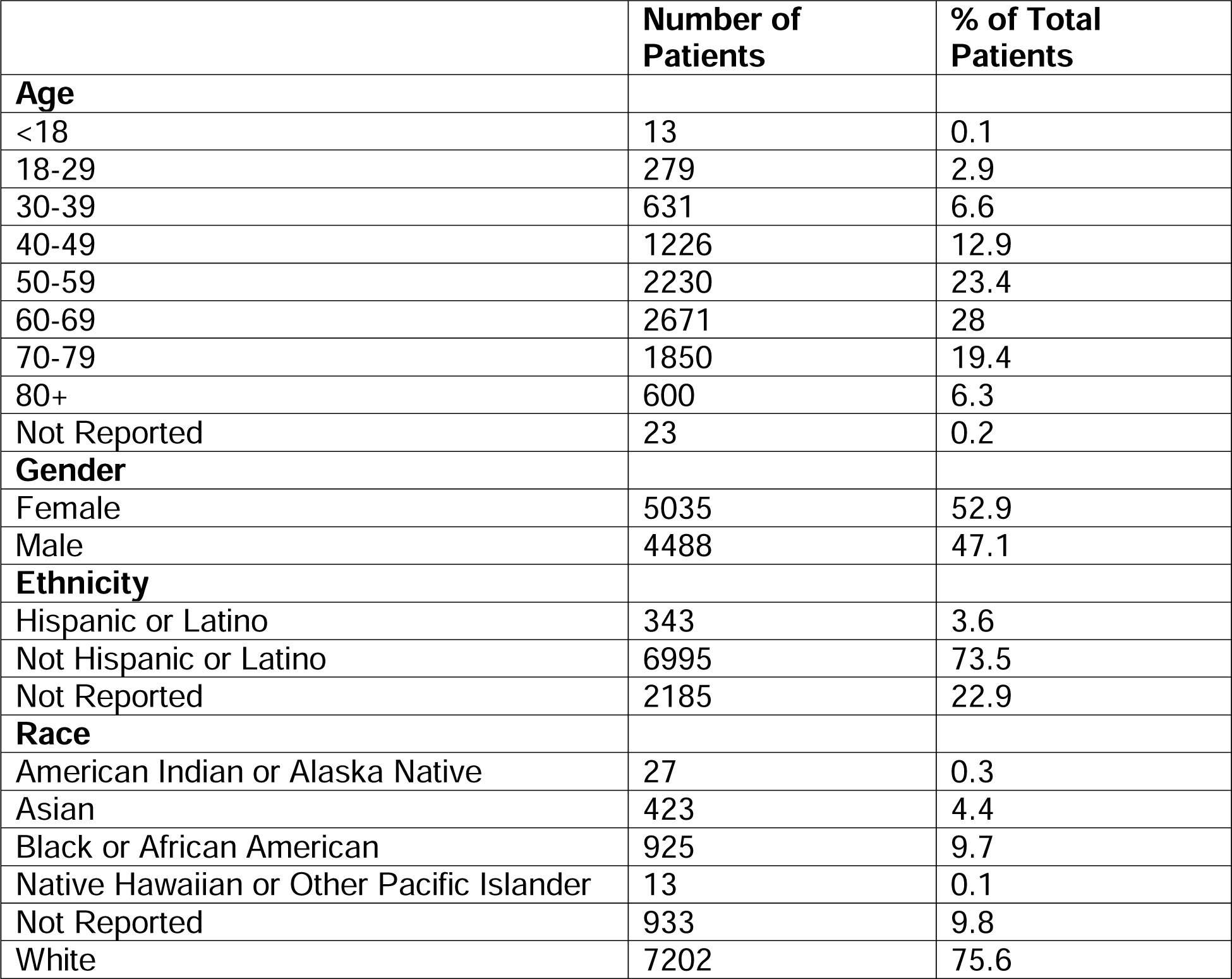
Demographic distribution of final pathology report dataset. See also Figure S5.

### Cancer Type Classification

As a proof-of-concept, we used the corpus to perform binary classification to predict cancer type (n=32). To utilize domain-relevant pretrained weights, we fine-tuned an existing model, ClinicalBERT.^26^ ClinicalBERT is a BERT-based model that had been trained on the clinical note set MIMIC-III^30^ and initialized with BioBERT^25^ weights. (BioBERT itself had been trained on PMC articles and Pubmed abstracts, and had been initialized with BERT-BASE.^24^) To prepare the text for input, we joined all lines and pages across each patient report. We partitioned the data into a train/validation/test split, stratifying by tissue type and holding out the test set until final evaluation.

Train/validation and test patient sets were consistent across demographic strata (Table S2). We trained in parallel 32 ClinicalBERT models^26^ for 10 epochs, across 10 random seeds per tissue type (see Methods). We identified models with maximal validation set AU-ROC and evaluated the performance of these models on the held-out test set. We were able to achieve an average test-set AU-ROC of 0.992 and average test-set AU- PRC of 0.903 (Fig. 3A-B).

**Figure 3.**
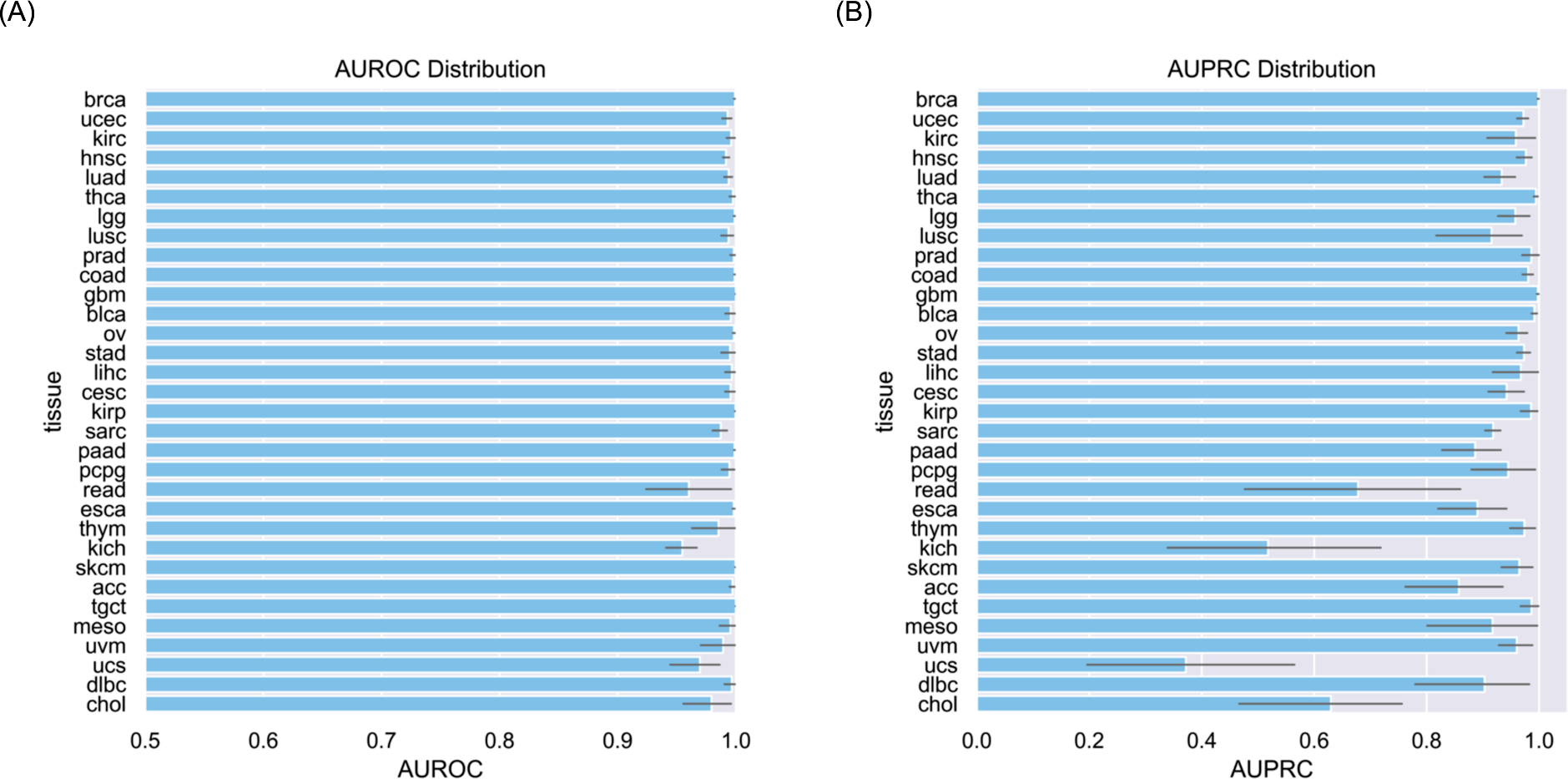
Model Performance for Proof-of-Concept Classification Task. Horizontal bar chart for (A) AU-ROC and (B) AU-PRC for test set performance of models trained across 10 random seeds, with 95% confidence interval. (A) and (B) are ordered by tissue prevalence, with higher-prevalence cancer types toward the top of each figure. See also Figure S4.

## Discussion

Pathology report text is generated routinely and ubiquitously across cancer care sites. In some medical centers, records can span decades, allowing for the research use of pathology reports in both retrospective and prospective analyses. Compared to whole slide image data, report text is substantially smaller in size and easier to work with. Text files require far less storage and model training requires much less run-time, especially important as memory and computing power can be cost-prohibitive. Reports reflect the expertise of practicing pathologists, who are typically equipped with years of specialty training and the morphological features they describe may prove helpful in training models to predict clinical targets.

The TCGA pathology report corpus can be utilized by researchers for a variety of analyses. For example, the text may be used as input for cancer subtype classification, survival prediction for increased prognostic accuracy, and information retrieval or named entity recognition (i.e., consistent extraction of specific information from report text). Directly, a clinical researcher could train and validate their model of interest on the TCGA corpus, and then apply that trained model to private patient data at their institution. This type of research could be performed for a specific cancer type or in a pan-cancer capacity. As models increase in capability, e.g. the recent advances in AI language models such as GPT4, the availability of relevant public text data will be essential for the benchmarking of relative model performance on pathology report text.

One of the main strengths of this dataset is that it is derived from the notes of many different pathologists at a wide range of institutions (Fig. 2D). This diversity will result in greater generalizability of models trained, particularly compared to models trained at single institutions. An additional benefit of this dataset is that it is already de-identified for public use and does not require specialized or controlled access, allowing its use as a convenient benchmark with which to compare different text-based models.

The TCGA pathology report corpus is enriched by additional patient data gathered by TCGA and accessible through its portal. These include histopathology slide imaging, clinical metadata, survival data, among other information (Table 2). The availability of these data opens the possibility of performing multimodal analyses, which may increase performance of downstream tasks.^31^ A limitation, however, of the TCGA dataset is that it does not contain clinical notes or symptom timelines, and the length of survival follow-up varies depending on cancer type.

**Table 2.** Selected Available Data for Patients in Final Pathology Report Set. Percent of patients with data availability, per cancer type. N = Number of patients, Eth. = Ethnicity, Prior Malig. = Prior Malignancy, OS = Overall Survival, PFI = Progression-Free Interval, DFI = Disease-Free Interval, derived from TCGA-CDR.28 All other columns derived from TCGA clinical and biospecimen metadata.^33^ Additional patient data, such as ICD-10 codes, sequencing data, transcriptomic data, and epigenetic data, are available through the TCGA portal.^33^

| Cancer Type | N | Age | Race | Eth. | Prior Malig. | Tumor Slides | Normal Slides | OS Events | PFI Events | DFI Events |
| --- | --- | --- | --- | --- | --- | --- | --- | --- | --- | --- |
| BRCA | 1034 | 100.00 | 90.81 | 83.17 | 99.90 | 100.00 | 15.18 | 14.02 | 13.54 | 7.93 |
| UCEC | 546 | 99.63 | 94.14 | 71.43 | 100.00 | 100.00 | 7.33 | 16.48 | 22.53 | 10.44 |
| KIRC | 525 | 100.00 | 98.67 | 71.05 | 100.00 | 100.00 | 82.29 | 33.52 | 29.71 | 2.29 |
| HNSC | 520 | 100.00 | 97.31 | 93.27 | 100.00 | 100.00 | 15.00 | 42.50 | 37.88 | 5.38 |
| LUAD | 488 | 97.95 | 88.93 | 77.46 | 100.00 | 100.00 | 42.01 | 36.68 | 41.19 | 17.83 |
| THCA | 487 | 100.00 | 81.72 | 79.88 | 100.00 | 100.00 | 19.92 | 3.29 | 10.06 | 5.13 |
| LGG | 469 | 100.00 | 97.87 | 93.18 | 100.00 | 100.00 | 0.00 | 23.67 | 34.33 | 3.41 |
| LUSC | 468 | 98.72 | 76.50 | 66.24 | 99.79 | 100.00 | 49.36 | 43.59 | 29.70 | 13.25 |
| PRAD | 446 | 100.00 | 97.09 | 80.72 | 100.00 | 100.00 | 26.01 | 1.79 | 16.14 | 4.93 |
| COAD | 418 | 100.00 | 58.61 | 56.46 | 100.00 | 99.76 | 21.05 | 19.86 | 27.03 | 5.50 |
| GBM | 399 | 100.00 | 96.99 | 82.21 | 7.77 | 100.00 | 1.25 | 75.69 | 79.95 | 0.50 |
| BLCA | 379 | 100.00 | 95.51 | 92.88 | 100.00 | 100.00 | 9.23 | 45.38 | 44.59 | 7.39 |
| OV | 371 | 100.00 | 93.26 | 47.17 | 2.70 | 100.00 | 19.95 | 52.02 | 65.77 | 30.73 |
| STAD | 361 | 99.17 | 84.21 | 70.36 | 100.00 | 100.00 | 25.76 | 40.44 | 31.58 | 8.86 |
| LIHC | 341 | 100.00 | 97.07 | 94.43 | 100.00 | 100.00 | 26.10 | 32.26 | 47.51 | 38.12 |
| CESC | 289 | 100.00 | 88.24 | 64.36 | 100.00 | 100.00 | 2.42 | 22.84 | 22.84 | 8.65 |
| KIRP | 280 | 99.29 | 95.00 | 87.50 | 100.00 | 100.00 | 30.00 | 13.93 | 19.29 | 8.93 |
| SARC | 249 | 100.0<br>0 | 96.39 | 87.55 | 100.0<br>0 | 100.0<br>0 | 8.43 | 37.75 | 53.01 | 24.90 |
| PAAD | 176 | 100.0<br>0 | 97.73 | 76.70 | 100.0<br>0 | 100.0<br>0 | 22.16 | 52.27 | 58.52 | 13.07 |
| PCPG | 174 | 100.0<br>0 | 97.70 | 80.46 | 100.0<br>0 | 100.0<br>0 | 2.87 | 2.87 | 10.92 | 2.30 |
| READ | 162 | 100.0<br>0 | 50.00 | 46.91 | 100.0<br>0 | 100.0<br>0 | 11.11 | 12.35 | 22.22 | 4.32 |
| ESCA | 146 | 100.0<br>0 | 86.30 | 38.36 | 100.0<br>0 | 100.0<br>0 | 44.52 | 48.63 | 45.89 | 3.42 |
| THYM | 114 | 100.0<br>0 | 98.25 | 87.72 | 100.0<br>0 | 100.0<br>0 | 7.02 | 5.26 | 16.67 | 0.00 |
| KICH | 112 | 100.0<br>0 | 98.21 | 66.07 | 100.0<br>0 | 100.0<br>0 | 62.50 | 10.71 | 14.29 | 5.36 |
| SKCM | 102 | 100.0<br>0 | 98.04 | 95.10 | 100.0<br>0 | 100.0<br>0 | 0.00 | 27.45 | 35.29 | 0.00 |
| ACC | 90 | 100.0<br>0 | 87.78 | 52.22 | 100.0<br>0 | 100.0<br>0 | 4.44 | 37.78 | 54.44 | 15.56 |
| TGCT | 87 | 100.0<br>0 | 94.25 | 88.51 | 100.0<br>0 | 100.0<br>0 | 0.00 | 2.30 | 18.39 | 13.79 |
| MESO | 79 | 100.0<br>0 | 100.0<br>0 | 87.34 | 100.0<br>0 | 100.0<br>0 | 1.27 | 83.54 | 72.15 | 8.86 |
| UVM | 65 | 100.0<br>0 | 61.54 | 60.00 | 100.0<br>0 | 100.0<br>0 | 0.00 | 26.15 | 33.85 | 0.00 |
| UCS | 56 | 100.0<br>0 | 98.21 | 76.79 | 100.0<br>0 | 100.0<br>0 | 10.71 | 62.50 | 66.07 | 17.86 |
| DLBC | 47 | 100.0<br>0 | 100.0<br>0 | 100.0<br>0 | 100.0<br>0 | 100.0<br>0 | 0.00 | 17.02 | 23.40 | 6.38 |
| CHOL | 43 | 100.0<br>0 | 97.67 | 93.02 | 100.0<br>0 | 100.0<br>0 | 39.53 | 48.84 | 51.16 | 20.93 |

The final text corpus presented here is moderately curated; data quality could be enhanced by applying additional cleaning steps for future analyses. For example, automated spelling correction could be applied in order to ensure that spelling mistakes made either in the original text or during OCR are corrected prior to model input tokenization. Depending on the model and tokenizer being applied, other pre- processing steps could include automated editing of punctuation or uncasing of the input text.

Finally, cancer-type classification was performed in this study as a proof-of-concept to illustrate the trainability of and information content within the corpus.

However, four cancer types (READ, KICH, UCS, CHOL) had relatively poor performance (low AU-PRC) for this classification task. Low AU-PRC may be a result of ClinicalBERT confusing one cancer type (e.g., UCS) with a similar cancer type (e.g., UCEC), particularly if the relative prevalence is severely imbalanced. Future work involving pathology reports for these low-prevalence cancer types could consider balancing classification^32^ or testing different models and tokenizers to potentially improve performance.

## Acknowledgments

Our work was supported by the following grant: R35GM131905.

## Data and code availability

https://github.com/tatonetti-lab/tcga-path-reports

## Methods

### TCGA Pathology Report Pre-Processing and Data Selection

We downloaded pathology reports, clinical metadata, and biospecimen metadata for all TCGA patients from the GDC portal.^33^ Each tumor sample has at most one associated pathology report (pathology_report_uuid), and each patient can have multiple samples. For case-based selection, we used sample.tsv (biospecimen directory). We removed patients with empty pathology_report_uuid values, removed patients with multiple pathology_report_uuid’s, and selected patients with “Primary Tumor” in the sample_type column. Next, we checked which patients matched with the TCGA Clinical Data Resource^28^ (a curated, comprehensive resource for TCGA outcomes data). We removed 72 patients either not found or found but lacking survival time within the TCGA-CDR.

For report-based filtering, we used OCR to identify reports for removal. We converted the reports from PDF-to-image and then image-to-text using pdf2image and pytesseract.^34–35^ We scanned the resultant text for key phrases for report exclusion. We removed 381 reports that contained the phrase “TCGA Missing Pathology Report Form” within any page (Fig S2A), 212 reports that contained the phrase “TCGA Pathologic Diagnosis Discrepancy Form” within any page (Fig S2B), and 14 reports of poor scan quality. Ultimately, 9,850 reports were selected for full text extraction and post-processing.

### Text Extraction and OCR Post-Processing

#### Text Extraction

Multiple OCR packages were tested for translation accuracy and output formatting consistency. A set of 50 randomly-chosen reports was used as a basis for comparison. First, we evaluated PyPDF2,^36^ a python package that converts PDF files directly to text. Although text translation performed reasonably well on the prototype set, there were a number of issues with the output files, including poorly translated TCGA quality control (QC) tables, incorrectly spaced words, and redaction bar artifacts in various sections of text. These factors made it infeasible to parse the output and achieve clean report text. Next, we evaluated the performance of pytesseract^35^ and Textract^29^ on the pathology report dataset. In order to use these packages, we performed a high-fidelity conversion of each page of the PDF prototype set to JPG image files using pdf2image.^34^ The python package pytesseract produced better quality text files in comparison with PyPDF2. The output text was largely structured the same as the input files, with no major word spacing issues. Barcodes and redaction bars were not translated at all, resulting in much cleaner output. However, pytesseract failed at handwriting translation, leading to mis-translated text in variable sections of each report that would be difficult to parse out in post-OCR processing.

Finally, we tested Textract, a software created by Amazon Web Services (AWS) that uses OCR and machine learning to convert images into text alongside structural annotation.^29^ In contrast to pytesseract and PyPDF2, output files include structural annotation in addition to text. For example, tables, selection elements, and hand-written lines are identified and annotated with bounding box coordinates within each report page. This feature is particularly helpful for parsing out mis-translated handwriting during post-processing and for filtering reports using table- or selection element-based filters (see Form Detection and Removal). In addition, we found that Textract produced cleaner output and consistently performed with higher translation accuracy on the prototype set (although, as with pytesseract, handwriting was not well-translated).

Based on these considerations, we selected Textract for use on the entire pathology report dataset.

For each report, page images were converted into byte arrays and processed on the AWS server. Due to AWS hard limits (<10k pixels/edge and <5MB total), we lowered the resolution slightly for 58 pages to conform. We converted 9,850 reports (25,478 pages) using the AnalyzeDocument function of Textract, with the table annotation option selected. We manually reviewed outlier short reports consisting of less than or equal to 5 lines of text (n=24 reports); we found that they contained clinically-relevant information and were therefore kept in the dataset.

#### Form Detection and Removal

Multiple-choice forms, consisting of questions with multiple-choice answer options were identified and removed from the dataset (Fig S2C). The multiple-choice selection elements were most frequently check-boxes, but the exact format varied. The forms themselves were variable in content (with disease-specific questions and answers), overall format, and number of selection elements per question. Because the selected option for each multiple-choice question is not detected by OCR, the resultant output text contains all multiple-choice options for each question. Some reports consisted entirely of multiple-choice forms and were fully removed, while others contained a mix of page types, in which case only form-containing pages were removed.

We first searched for reports that potentially contained multiple-choice content. As an initial filter, we selected reports based on structural elements, including the total number of tables per report, the total number of selection elements per report, and the average and maximum number of selection elements per page. All structural elements considered in this section were annotated by Textract, with annotation data represented by the BlockType attribute of each page response block. We employed various empirically-derived thresholds for this initial filter, starting with the clearest outliers and then including medium outliers, finding additional form reports in the medium outlier set upon manual review. However, we found that this first-level filter was not specific enough, including many non-forms in the selected report sets. We also observed that only a few cancer types had form-style pathology reports in this dataset.

We therefore added a second filter consisting of custom disease-specific keywords, based on manual review of a subset of reports selected from the structural elements filter. Keywords were drawn from both question and answer text, with a preference for unique phrases that were unlikely to appear elsewhere in standard report text. The number of matched keywords required for report selection was adjusted depending on the results for each disease filter. For example, colon cancer pathology reports were identified as likely forms if they contained at least 2 of the following keywords: “Signet Ring Feature:”, “Histologic Heterogeneity:”, “Crohn’s like reaction”, “Plasma cell rich stroma”, “Angiolymphatic Invasion:”, “Garland Necrosis present:”, “TIL Cells / HPF”, “Pathologist Comment:”. As another example, liver-related forms were identified by 14 keywords, including “Hepatitis (specify type)”, and “(check all that apply)”, and cervix-related forms were identified using 21 keywords across multiple pages. We additionally incorporated fuzzy-matching to the keyword filter to account for misspelled text (either misspelled via OCR translation error or within the original pathology report text).

Although the keyword filter greatly increased specificity, enriching report sets for form content, the final filtered report sets were not perfectly specific. We therefore manually reviewed all reports that passed the structural element and keyword-matching filters to ensure we ultimately removed only form-reports from the overall dataset. After form removal, 9,547 reports (24,214 pages) remained.

#### Table Detection and Removal

We used 9 section-header keywords to identify TCGA QC tables within each report (Fig S3). The section headers were largely typed text, free of handwriting annotation, and Textract transcribed these keywords sufficiently for fuzzy detection. Fuzzy-matching error allowance varied according to the observed frequency of mis- translation for each keyword. The relative location of the section headers within each table was consistent across reports. As such, we drew a custom bounding box for each keyword detected, and then merged the keyword-based bounding boxes to form a “maximum bounding box” around the entire detected table.

To check that this table detection method performs accurately across the overall dataset, we probed the results in a number of ways. First, we manually scored a prototype set of 50 randomly-selected reports, finding that all detected tables were true tables, and all true tables were detected (no false negatives or false positives). Next, we tested whether a single matched keyword was sufficient to distinguish table content from main text. Checking for false-positives, we manually reviewed all reports for which only one keyword was fuzzy-matched. Upon reviewing 115 pages that met this criterion, we found that all detected bounding boxes were true QC tables. This aligns with our observation that the terms used in QC table section headers are distinct from the general vocabulary used in the main text. We also examined large max bounding box outliers to confirm that main report text was not fuzzy-matched by our table detection method. We found that these reports (n=39) had reasonably-sized max bounding boxes, and the bounding boxes did not overlap with any clinically-relevant, non-QC-table lines.

Once the tables had been detected, we removed them by removing any lines overlapping with the max bounding box. We set an overlap threshold, or the minimum area overlap between the bounding box of a given line and the max bounding box of a QC table for the line to be considered as part of the table. A smaller overlap threshold would include lines that were further from the table, as less overlap area would be required for the line to be considered part of the table. To determine the appropriate overlap threshold, we assembled a randomly-selected subset (n=4000 pages) and manually examined pages containing lines within specified overlap thresholds. Between thresholds .35-.25, no clinically-relevant, non-table-related lines were selected; however for lines with overlap <= .25, some clinically-relevant, non-table-related lines were captured. We therefore moved forward with a minimum .25 area overlap threshold, and removed all QC-table-related lines from the dataset.

#### Handwriting and Keyword Removal

We implemented additional filters to clean the text before dataset finalization.

First, we sought to remove TCGA handwritten annotations; these were typically incorrectly OCR-translated, and not part of standard pathology report text (i.e., an artifact of the TCGA data collection process). We selected for lines that consisted entirely of Textract-annotated handwritten words, removing approximately 120,000 lines in this step. In addition, we sought to remove any clinically-irrelevant TCGA identification data or site-specific text (such as clinic-specific section headers), with the goal of reducing any potentially confounding elements within the text itself. We manually reviewed 500 randomly-selected report pages and compiled a list of 312 regular expressions. Approximately 100,000 additional lines were removed at this stage. After joining all lines with period delimiters and joining report pages, the final dataset consisted of 9,523 reports (23,909 pages) across 32 cancer types.

### Cancer Type Classification

We performed binary cancer type classification by fine-tuning Bio+Clinical BERT^26^ and using TCGA project_id as the prediction target. We trained each model in parallel, with 32 separate cancer type experiments. We split the data into train/validation/test sets, stratifying by cancer type. To establish confidence intervals for model performance, we ran 10 different random seeds for each experiment, resulting in 320 models trained and evaluated. We trained the models with default parameters, except for the following: per_device_train_batch_size set to 16 (for smoother training curves and reduced run-time); AU-ROC was used for performance evaluation; models were saved and evaluated every 32 steps (more often than default). Model input was truncated at 512 tokens per patient report, which is the maximum number of input tokens that ClinicalBERT is able to utilize. For evaluation, we applied a softmax on raw model scores, and used the transformed values for ROC and PR curve construction.

We trained all cancer-type models across 10 random seeds for 10 epochs. Each model used approximately 7,620 seconds of run-time, for a total training time of 11.3 days. The best models, as determined by highest validation set AU-ROC, were then applied to the test set for evaluation. AU-ROC was consistently high across cancer types, with narrow confidence intervals (Fig 3A). AU-PRC was more variable across cancer types and exhibited wider confidence intervals (Fig 3B). Performance amongst tissues with lower prevalence was generally worse as compared with tissues of higher prevalence. This is to be expected, as models trained on limited sample size are presented with fewer examples with which to learn their intended classification target, and generally benefit from larger sample sizes with greater total information content.

Individual per-tissue ROC and PR curves were plotted for comparison (Fig S4).

## Supplemental Titles and Legends

Figure S1. Related to Figure 1 and Methods (“Text Extraction and OCR Post-Processing”). Report Examples – TCGA-Inserted Within-Report Metadata Artifacts.

(A) Redaction Bars (B) TCGA Barcode (C) TCGA QC Table and Handwritten Annotation.

Figure S2. Related to Methods (“Form Detection and Removal”). Report Examples – Removed Forms. (A) TCGA Missing Pathology Report Form (B) TCGA Pathologic Diagnosis Discrepancy Form (C) Additional Multiple-Choice Forms, demonstrating variability.

Figure S3. Related to Figure 1 and Methods (“Table Detection and Removal”). TCGA QC Table – Automated Max Bounding Box Detection Example.

Figure S4. Related to Figure 3. Average ROC (A) and PR (B) curves for all tissue models (10 epochs, 10 random seeds, test set performance). Plots are ordered according to descending prevalence within the final post-processed dataset.

Figure S5. Related to **Table 1**. Additional Characteristics of Final Dataset. (A) Pages per Report, per-Tissue Distribution. (B) Tissue Sites (Institutions) per-Tissue Distribution. (C) Age Distribution. (D) Race Distribution. (E) Gender Distribution.

Table S1. Patients per cancer type in final dataset. Related to Figure 1.

Table S2. Demographic table across train and test sets. Related to Table 1 and Methods (“Cancer Type Classification”).

