## Supplemental Data 1 for "Benchmark Pathology Report Text Corpus with Cancer Type Classification"

(A)

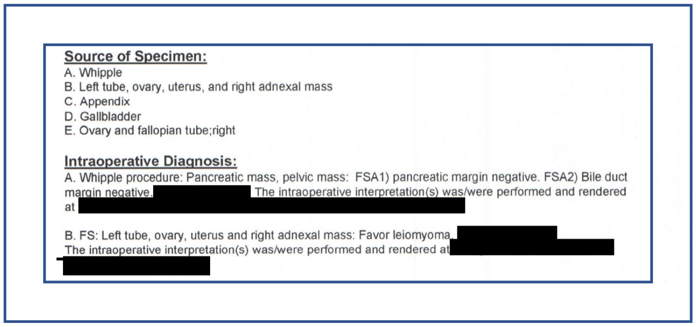

**Source of Specimen:**
A. Whipple
B. Left tube, ovary, uterus, and right adnexal mass
C. Appendix
D. Gallbladder
E. Ovary and fallopian tube;right

**Intraoperative Diagnosis:**
A. Whipple procedure: Pancreatic mass, pelvic mass: FSA1) pancreatic margin negative. FSA2) Bile duct margin negative. [redacted] The intraoperative interpretation(s) was/were performed and rendered at [redacted]

B. FS: Left tube, ovary, uterus and right adnexal mass: Favor leiomyoma [redacted]
The intraoperative interpretation(s) was/were performed and rendered at [redacted]

(B)

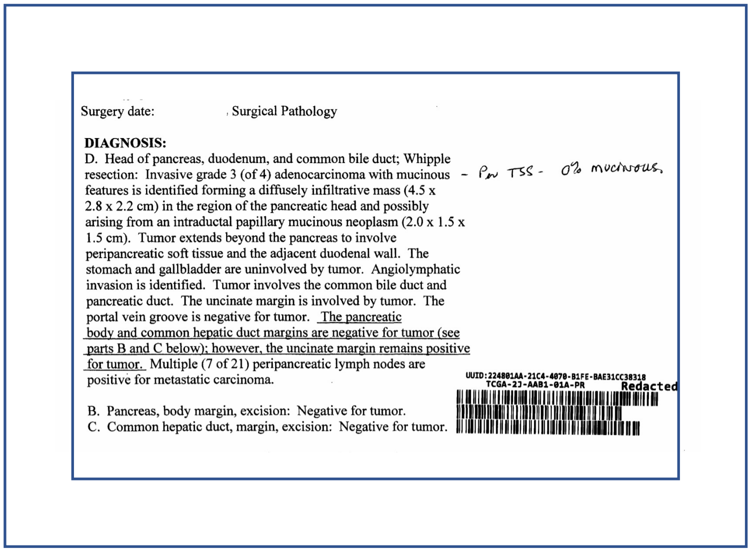

Surgery date: Surgical Pathology

**DIAGNOSIS:**
D. Head of pancreas, duodenum, and common bile duct; Whipple resection: Invasive grade 3 (of 4) adenocarcinoma with mucinous features is identified forming a diffusely infiltrative mass (4.5 x 2.8 x 2.2 cm) in the region of the pancreatic head and possibly arising from an intraductal papillary mucinous neoplasm (2.0 x 1.5 x 1.5 cm). Tumor extends beyond the pancreas to involve peripancreatic soft tissue and the adjacent duodenal wall. The stomach and gallbladder are uninvolved by tumor. Angiolymphatic invasion is identified. Tumor involves the common bile duct and pancreatic duct. The uncinate margin is involved by tumor. The portal vein groove is negative for tumor. The pancreatic body and common hepatic duct margins are negative for tumor (see parts B and C below); however, the uncinate margin remains positive for tumor. Multiple (7 of 21) peripancreatic lymph nodes are positive for metastatic carcinoma.

- Per TSS - 0% mucinous.

B. Pancreas, body margin, excision: Negative for tumor.
C. Common hepatic duct, margin, excision: Negative for tumor.

UUID:224801AA-21C4-4070-B1FE-BAE31CC3B318
TCGA-2J-AAB1-01A-PR
Redacted

(C)

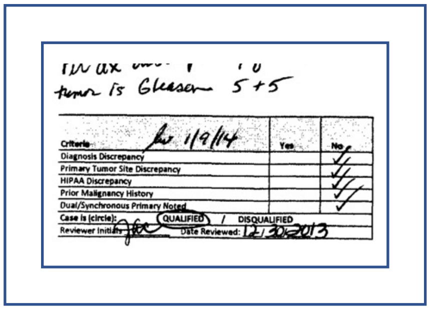

tumor is Gleason 5+5

| Criteria | Yes | No |
|---|---|---|
| Diagnosis Discrepancy | | ✓ |
| Primary Tumor Site Discrepancy | | ✓ |
| HIPAA Discrepancy | | ✓ |
| Prior Malignancy History | | ✓ |
| Dual/Synchronous Primary Noted | | ✓ |
| Case is (circle): QUALIFIED / DISQUALIFIED | | |
| Reviewer Initials: [illegible] Date Reviewed: 12/30/2013 | | |

1/9/14

**Figure S1.** Related to Figure 1 and Methods ("Text Extraction and OCR Post-Processing").
**Report Examples – TCGA-Inserted Within-Report Metadata Artifacts.**
(A) Redaction Bars (B) TCGA Barcode (C) TCGA QC Table and Handwritten Annotation.

(A)

Page 1 V3.00 110614

**TCGA Missing Pathology Report Form**

**Instructions:** *The TCGA Missing Pathology Report Form should be completed for cases for which a pathology report is not available.*

UUID:79806E48-15D0-45F9-860D-182787298EBA
TCGA-25-1628-01A-PR Redacted

Completed Date (MM/DD/YYYY): 12 / 29 / 2014

**General Pathology Report Information**

| # | Data Element | Entry Alternatives | Working Instructions |
|---|---|---|---|
| 1 | Tumor type: | OV | Provide the tumor type of the case. |
| 2 | BCR specimen originally sent to: | ☐ NCH<br>☑ IGC | Indicate to which BCR location the case was originally sent. |
| 3 | Date specimen received at BCR: | 5 / 13 / 2009 | Provide the date (MM/DD/YYYY) of shipment arrival at the Biospecimen Core Resource (BCR). |
| 4 | ICD-O-3 Histology Code:<br>- For Specimen: (CQCF)<br>- For Case: (patient diagnosis, if available elsewhere) | 8441/3<br>______ | Provide the histology code for the sample from the Case Quality Control Form (CQCF) and the overall case (patient diagnosis, if different). |
| 5 | ICD-O-3 Site Code:<br>- For Specimen: (CQCF)<br>- For Case: (patient diagnosis, if available elsewhere) | C56.9<br>______ | Provide the site code for the sample from the Case Quality Control Form (CQCF) and the overall case (patient diagnosis, if different). |

(B)

**TCGA Pathologic Diagnosis Discrepancy Form** V4.00

Instructions: *The TCGA Pathologic Diagnosis Discrepancy Form should be completed when the pathologic diagnosis documented on the initial pathology report for a case submitted for TCGA is inconsistent with the diagnosis provided on the Case Quality Control Form completed for the submitted case.*

Tissue Source Site (TSS): ___ _TSS Identifier: ________ TSS Unique Patient Identifier: _

Completed By *(Interviewer Name on OpenClinica)*: _ ____Completed Date: _ __________

Diagnosis Information

| # | Data Element | Entry Alternatives | Working Instructions |
|---|---|---|---|
| 1 | Pathologic Diagnosis Provided on Initial Pathology Report | 80% epithelioid cells<br>20% spindle cells | Provide the diagnosis/ histologic subtype(s) documented on the initial pathology report for this case. If the histology for this case is mixed, provide all listed subtypes. |
| 2 | Histologic features of the sample provided for TCGA, as reflected on the CQCF. | 61-90% epithelioid cells<br>1-30% spindle cells | Provide the histologic features selected on the TCGA Case Quality Control Form completed for this case. |

Discrepancy between Pathology Report and Case Quality Control Form

| # | Data Element | Entry Alternatives | Working Instructions |
|---|---|---|---|
| 3 | Provide the reason for the discrepancy between the pathology report and the TCGA Case Quality Control Form. | The section of the frozen sample used for sending to TCGA corresponds to a section of the tumor where the epithelioid part is predominant.<br>The percentages mentioned on the initial Pathology Report correspond to an average on the whole embedded sections used for establish the Pathology Report. | Provide a reason describing why the diagnosis on the initial pathology report for this case is not consistent with the diagnosis selected on the TCGA Case Quality Control Form. |
| 4 | Name of TSS Reviewing Pathologist or Biorepository Director | | Provide the name of the pathologist who reviewed this case for TCGA. |

(C)

**Consolidated Pathology Diagnosis**

**Histological Pattern**

| Cell Distribution | + | - | Structural Pattern | + | - |
|---|---|---|---|---|---|
| Diffuse | | ✓ | Streaming | | |
| Mosaic | ✓ | | Storiform | | |
| Necrosis | | ✓ | Fibrosis | | |
| Lymphocytic Infiltration | ✓ | | Palisading | | |
| Vascular Invasion | | ✓ | Cystic Degeneration | | |
| Clusterized | | ✓ | Bleeding | | |
| Alveolar Formation | | ✓ | Myxoid Change | | |
| Indian File | | ✓ | Psammoma/Calcification | | |

**Cellular Differentiation**

| Squamous | + | - | Adenomatous | + | - | Sarcomatous | + | - | Lymphomatous | + | - |
|---|---|---|---|---|---|---|---|---|---|---|---|
| Squamoid Cell | | | Glandular cell | ✓ | | Round Cell | | | Large Cell | | |
| Spindle Cell | | | Cell Stratification | ✓ | | Fibroblast | | | Small Cell | | |
| Keratin | | | Secretion | ✓ | | Osteoblast | | | RS Cell/RS Like | | |
| Desmosome | | | Intracyt. Vacuole | ✓ | | Lipoblast | | | Inflam. Cell | | |
| Pearl | | | Gland formation | | ✓ | Myoblast | | | Plasma Cell | | |

**Cellular Differentiation:** ☒ Well ☐ Moderate ☐ Poor

**Nuclear Appearance**

| Nuclear Atypia: | 0 | I | II | III |
|---|---|---|---|---|
| Aniso Nucleosis | | ✓ | | |
| Hyperchromatism | | ✓ | | |
| Nucleolar Prominent | | ✓ | | |
| Multinucleated Giant Cell | | ✓ | | |
| Mitotic Activity | | ✓ | | |
| **Nuclear Grade:** | | ✓ | | |

D1 80% D2 80% D3 80% D4 80%

**Final Pathology Report**

**Histological Diagnosis:** Hepatocellular Carcinoma Trabecular type **Grade:** 1

**Comments:**

SYNOPTIC – PRIMARY LUNG TUMORS [redacted]

A. Location: 1
1. RUL 3. RLL 5. LUL
2. RML 4. LLL 6. Bronchus intermedius
B. Procedure: 2
1. Wedge/Segmental 3. Bilobectomy
2. Lobectomy 4. Pneumonectomy
C. Size of tumor (maximum dimension): 4.2 cm.
D. Satellite nodules: 2
1. yes 2. No
D1. If yes, Where: #
1. Same lobe 2. Psilateral lobes 3. Central lobe
E. Type: 3
1. Invasive squamous carcinoma 11. Fetal type adenocarcinoma
2. Basaloid squamous carcinoma 12. Small cell carcinoma
3. Invasive adenocarcinoma 13. Carcinoid
4. Bronchioloalveolar carcinoma, nonmucinous type 14. Atypical carcinoid
5. Bronchioloalveolar carcinoma, mucinous type 15. Sarcoma
6. Large cell carcinoma 16. Adenosquamous carcinoma
7. Clear cell carcinoma 17. Mixed non-small cell carcinoma, NOS
8. Giant cell carcinoma 18. Carcinosarcoma
9. Sarcomatoid carcinoma 19. Other
10. Blastoma
F. Architectural grade: 1
1. well 2. moderate 3. poor/undifferentiated
G. Nuclear grade: 1
1. Low 2. Intermediate 3. High
H. Central vs. peripheral origin: 1
1. yes 2. no
I. Visceral pleural invasion through elastica: 2
1. yes 2. no
J. Parietal pleural invasion: 2
1. yes 2. no

ICD-O-3
Carcinoma, Squamous cell NOS 8070/3
Site: (R) Lung, upper & lower lobes C34.8
gW 2/1/13

Synoptic translated report

UUID:81068415-BBAA-432E-97C2-14E68559D41A
TCGA-NC-A5HT-01A-PR Redacted

**Site** : Right lung superior and middle lobes
☒ Central ☐ Peripheral ☐ NA

**Number of lesion** :
1. Lung squamous cell carcinoma

**Tumor size:**
1. 4.7 cm (diameter)

**Visceral pleural invasion:** ☐Yes ☒No ☐NA

**Chest wall invasion** : ☐Yes ☒No ☐NA

SYNOPTIC - PRIMARY INVASIVE CARCINOMA OF BREAST

A. Laterality: 2
1. Right 2. Left
B. Procedure: 2
1. Segmentectomy 3. Modified radical mastectomy
2. Simple mastectomy 4. Re-excision
C. Location: 4, 2
1. Central subareolar 4. LOQ
2. UOQ 5. LIQ
3. UIQ 6. Not specified
D. Size of tumor (maximum dimension invasive component by gross or microscopic exam): 2.6 and 3.0 cm
E. Type (invasive component): 1
1. Ductal, NOS 5. Cribriform 9. Metaplastic
2. Tubular 6. Papillary 10. Other
3. Mucinous 7. Lobular
4. Medullary 8. Carcinoid like
F. If lobular carcinoma, specify type: N/A
1: Classical 4. Signet ring 7. Pleomorphic
2. Solid 5. Trabecular
3. Alveolar 6. Tubulobular
G. Nottingham Score:
G1. Nuclear grade: 3
G2. Tubule formation: 3
G3. Mitotic activity score: 3
G4. Total Nottingham score: 9
G5. Nottingham grade (1, 2, 3): #
H. Angiolymphatic invasion: 2
1. No 2. Yes
I. Dermal lymphatic invasion: 2
1. Yes 2. No 3. Not applicable
J. Calcification: 1
1. No 2. Yes – benign zones 3. Yes – malignant zones
K. Type of in situ component: 2
1. Cribriform 4. Micropapillary 7. Lobular
2. Solid 5. Apocrine
3. Papillary 6. Comedo
L. Percentage of tumor occupied by in situ component: ≤5%
M. Surgical margins involved by invasive component: 1
1. No 2. Yes – focal 3. Yes – diffuse
N. Surgical margins involved by in situ component: 1
1. No 2. Yes – focal 3. Yes - diffuse

**Path (First Tumor)**

| | | | | |
|---|---|---|---|---|
| Tumor Site: | Descending Colon | | | |
| Date of Cancer Sample Procurement: | | | | |
| Histology: | Adenocarcinoma | | | |
| Description of other histology: | | | | |
| Grade: | Moderately Differentiated | | | |
| Mucinous: | ○ No | ○ Yes | ● Yes (Focal) | ○ Unknow |
| Signet Ring Feature: | ● No | ○ Yes | ○ Yes (Focal) | ○ Unknow |
| Histologic Heterogeneity: | ○ No | ● Yes | ○ Unknown | |
| Host Response: | None | | | |
| Crohn's like reaction | ● None | ○ Yes | ○ Unknown | |
| Plasma cell rich stroma | ○ No | ○ Yes | ○ Unknown | |
| Growth Pattern: | ● Expansile | ○ Invasive | ○ Expansile and Invasive | ○ Unknow |
| Inflammatory Bowel Disease | ● No | ○ Yes | ○ Unknown | |

**Figure S2.** Related to Methods ("Form Detection and Removal"). **Report Examples – Removed Forms.** (A) TCGA Missing Pathology Report Form (B) TCGA Pathologic Diagnosis Discrepancy Form (C) Additional Multiple-Choice Forms, demonstrating variability.

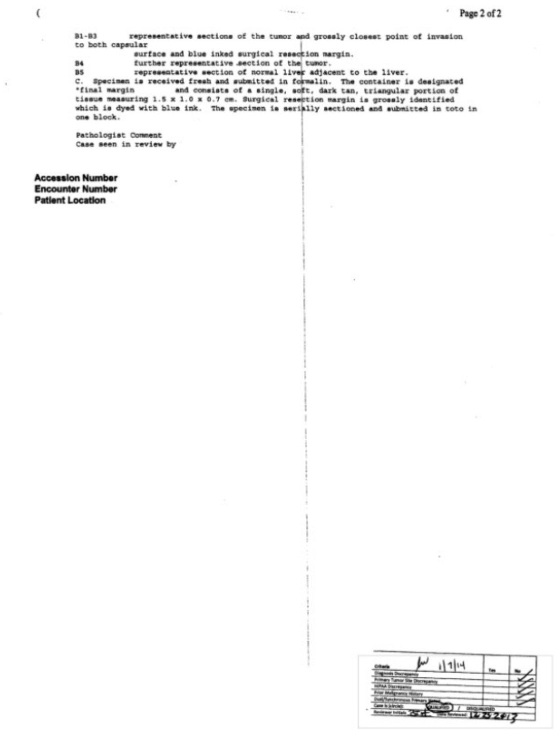

( Page 2 of 2

B1-B3 representative sections of the tumor and grossly closest point of invasion
to both capsular
surface and blue inked surgical resection margin.
B4 further representative section of the tumor.
B5 representative section of normal liver adjacent to the liver.
C. Specimen is received fresh and submitted in formalin. The container is designated
"final margin and consists of a single, soft, dark tan, triangular portion of
tissue measuring 1.5 x 1.0 x 0.7 cm. Surgical resection margin is grossly identified
which is dyed with blue ink. The specimen is serially sectioned and submitted in toto in
one block.

Pathologist Comment
Case seen in review by

**Accession Number**
**Encounter Number**
**Patient Location**

| Criteria | Yes | No |
|---|---|---|
| Diagnosis Discrepancy | | ✓ |
| Primary Tumor Site Discrepancy | | ✓ |
| HIPAA Discrepancy | | ✓ |
| Prior Malignancy History | | ✓ |
| Dual/Synchronous Primary Noted | | ✓ |
| Case is (circle): QUALIFIED / DISQUALIFIED | | |
| Reviewer Initials: [illegible] Date Reviewed: [illegible] | | |

**Figure S3.** Related to Figure 1 and Methods ("Table Detection and Removal").
**TCGA QC Table – Automated Max Bounding Box Detection Example.**

(A)

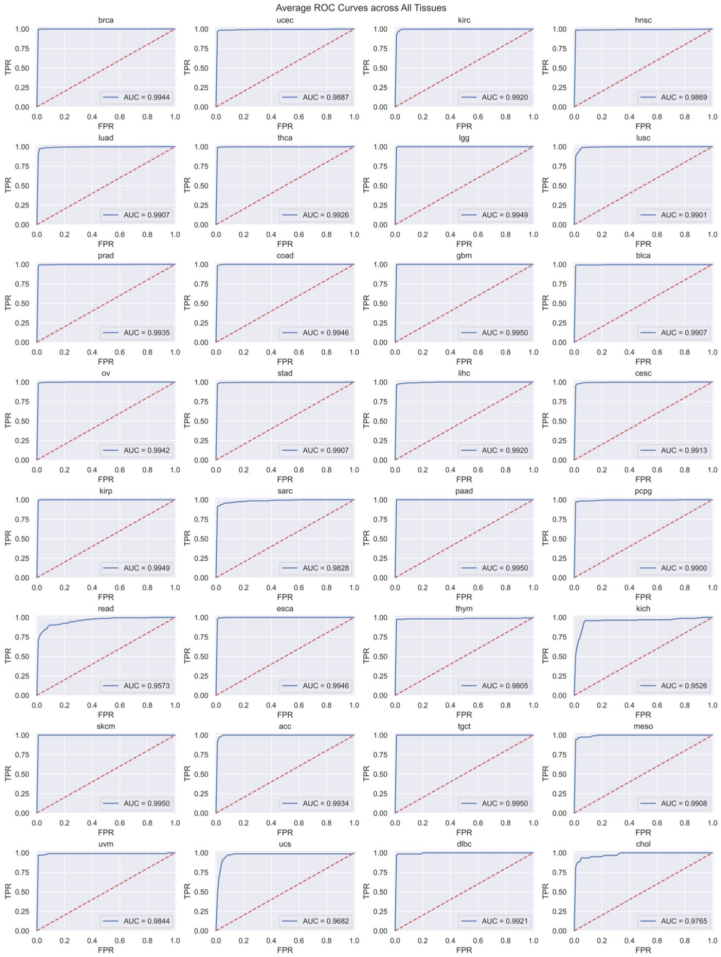


(B)

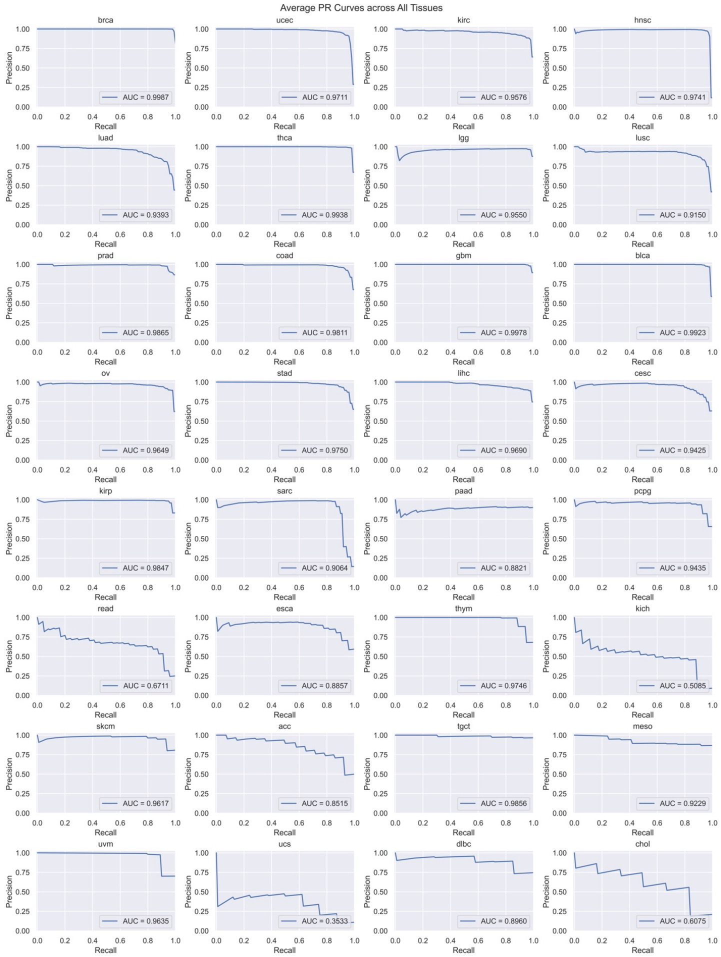


**Figure S4.** Related to Figure 3. Average ROC (A) and PR (B) curves for all tissue models (10 epochs, 10 random seeds, test set performance). Plots are ordered according to descending prevalence within the final post-processed dataset.

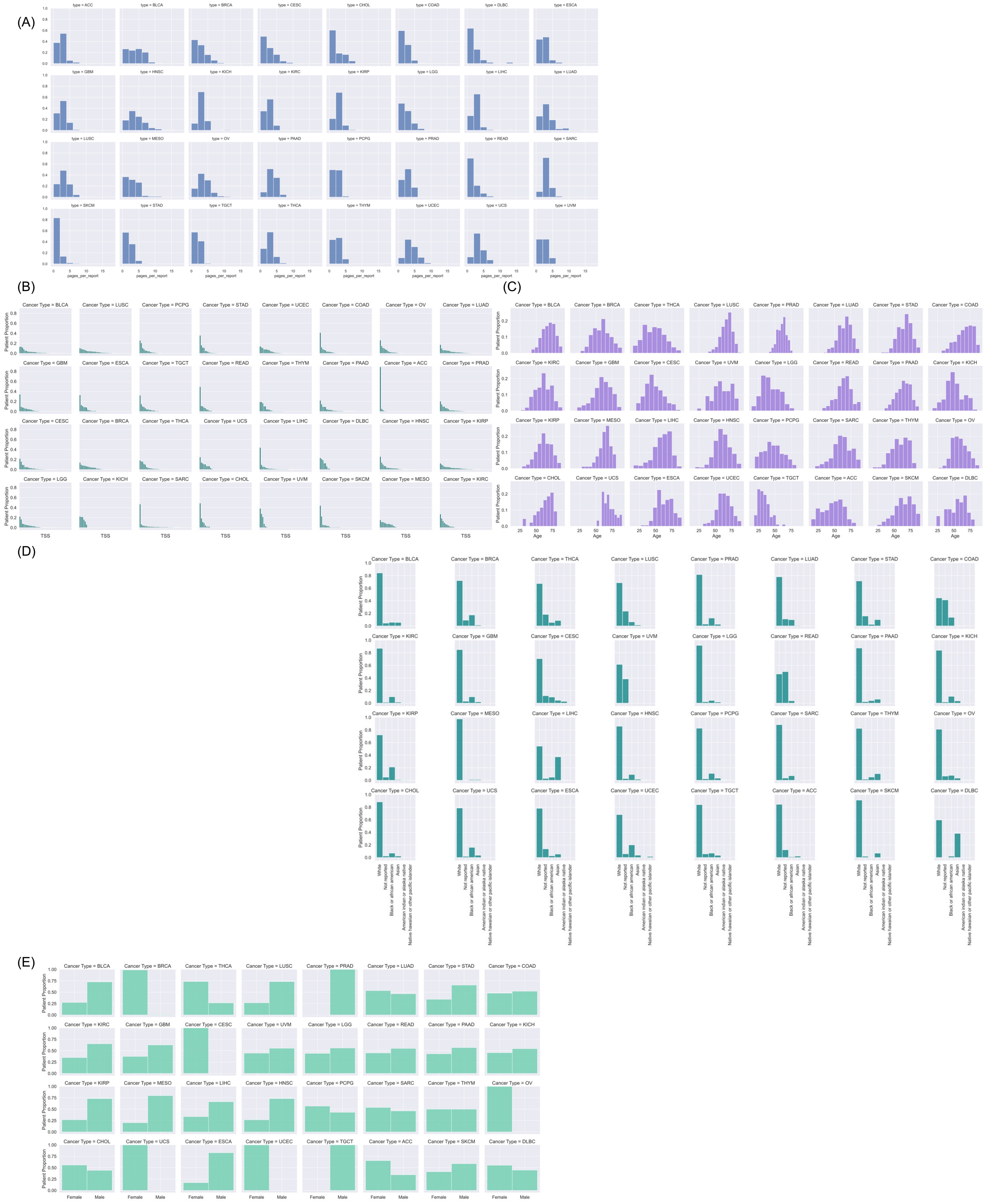


**Figure S5.** Related to Table 1. **Additional Characteristics of Final Dataset.** (A) Pages per Report, per-Tissue Distribution. (B) Tissue Sites (Institutions) per-Tissue Distribution. (C) Age Distribution. (D) Race Distribution. (E) Gender Distribution.
