## Supplemental Data 2 for "Benchmark Pathology Report Text Corpus with Cancer Type Classification"

**Table S1.** Patients per cancer type in final dataset. Related to Figure 1.

| Cancer Type | Number Patients Removed | Percent Patients Removed | Number Patients Remaining | Percent Patients Remaining |
|---|---|---|---|---|
| BRCA | 63 | 5.74 | 1034 | 94.26 |
| UCEC | 2 | 0.36 | 546 | 99.64 |
| KIRC | 12 | 2.23 | 525 | 97.77 |
| HNSC | 8 | 1.52 | 520 | 98.48 |
| LUAD | 34 | 6.51 | 488 | 93.49 |
| THCA | 20 | 3.94 | 487 | 96.06 |
| LGG | 46 | 8.93 | 469 | 91.07 |
| LUSC | 36 | 7.14 | 468 | 92.86 |
| PRAD | 54 | 10.8 | 446 | 89.2 |
| COAD | 41 | 8.93 | 418 | 91.07 |
| GBM | 196 | 32.94 | 399 | 67.06 |
| BLCA | 33 | 8.01 | 379 | 91.99 |
| OV | 216 | 36.8 | 371 | 63.2 |
| STAD | 82 | 18.51 | 361 | 81.49 |
| LIHC | 36 | 9.55 | 341 | 90.45 |
| CESC | 18 | 5.86 | 289 | 94.14 |
| KIRP | 11 | 3.78 | 280 | 96.22 |
| SARC | 12 | 4.6 | 249 | 95.4 |
| PAAD | 9 | 4.86 | 176 | 95.14 |
| PCPG | 5 | 2.79 | 174 | 97.21 |
| READ | 8 | 4.71 | 162 | 95.29 |
| ESCA | 39 | 21.08 | 146 | 78.92 |
| THYM | 10 | 8.06 | 114 | 91.94 |
| KICH | 1 | 0.88 | 112 | 99.12 |
| SKCM | 368 | 78.3 | 102 | 21.7 |
| ACC | 2 | 2.17 | 90 | 97.83 |
| TGCT | 47 | 35.07 | 87 | 64.93 |
| MESO | 8 | 9.2 | 79 | 90.8 |
| UVM | 15 | 18.75 | 65 | 81.25 |
| UCS | 1 | 1.75 | 56 | 98.25 |
| DLBC | 1 | 2.08 | 47 | 97.92 |
| CHOL | 2 | 4.44 | 43 | 95.56 |

**Table S2.** Demographic table across train and test sets. Related to Table 1 and Methods ("Cancer Type Classification").

| | N (Train) | P (Train) | N (Test) | P (Test) |
|---|---|---|---|---|
| **Age** | | | | |
| 0-18 | 12 | 0.1 | 1 | 0.1 |
| 18-29 | 236 | 2.9 | 43 | 3 |
| 30-39 | 540 | 6.7 | 91 | 6.4 |
| 40-49 | 1043 | 12.9 | 183 | 12.8 |
| 50-59 | 1881 | 23.2 | 349 | 24.4 |
| 60-69 | 2288 | 28.3 | 383 | 26.8 |
| 70-79 | 1575 | 19.5 | 275 | 19.2 |
| 80+ | 501 | 6.2 | 99 | 6.9 |
| Not Reported | 18 | 0.2 | 5 | 0.3 |
| **Gender** | | | | |
| Female | 4243 | 52.4 | 792 | 55.4 |
| Male | 3851 | 47.6 | 637 | 44.6 |
| **Ethnicity** | | | | |
| Hispanic or Latino | 282 | 3.5 | 61 | 4.3 |
| Not Hispanic or Latino | 5972 | 73.8 | 1023 | 71.6 |
| Not Reported | 1840 | 22.7 | 345 | 24.1 |
| **Race** | | | | |
| American Indian or Alaska Native | 24 | 0.3 | 3 | 0.2 |
| Asian | 355 | 4.4 | 68 | 4.8 |
| Black or African American | 782 | 9.7 | 143 | 10 |
| Native Hawaiian or Other Pacific Islander | 12 | 0.1 | 1 | 0.1 |
| Not Reported | 771 | 9.5 | 162 | 11.3 |
| White | 6150 | 76 | 1052 | 73.6 |
